# Portable Objective Diagnostics using Visual Evoked Potentials for Age-related Macular Degeneration

**DOI:** 10.1101/2020.01.03.20016451

**Authors:** Craig Versek, S. Mohammad Ali Banijamali, Peter J. Bex, Kameran Lashkari, Sagar V. Kamarthi, Srinivas Sridhar

## Abstract

Delayed Dark Adapted vision Recovery (DAR) is a biomarker for Age-related Macular Degeneration (AMD), however its measurement is burdensome for patients and examiners. We developed a portable, wireless, quick-setup system that employs a headset with a smartphone to deliver controlled photobleach and monocular pattern reversal stimuli, while using custom electroencephalography (EEG) electrodes and electronics in order to measure objective Dark Adapted Visual Evoked Potentials (DAVEP) at multiple locations of the visual field - all in one comfortable 20-minute session, without requiring subject reporting. DAVEP responses post photobleach (up to 15 minutes), were measured concurrently in both eyes of 13 patients with AMD and 8 others not diagnosed with AMD. New, unexpected features at high latencies were observed in the DAVEP responses to scotopic stimulus intensities. The amplitude recovery of the DAVEP response was significantly delayed in AMD patients compared with controls. We developed DAVEP1 scores, a simple metric for DAR, using it to successfully identify all 100% of AMD subjects and classify 90% of subject eyes correctly. Deficits in DAR in patients with AMD can be identified using this objective VEP based system and the DAVEP1 metric, a promising new objective biomarker for this disease that can be easily tested in a clinic.

## Introduction

Age-related macular degeneration (AMD) causes progressive loss of central vision and is the leading cause of irreversible blindness in the developed world [1]. The estimated number of people with AMD is approximately 2.1M in the US [2], and the disease is expected to affect 20-25 million people worldwide by 2030 [3]. AMD causes vision loss in the central retinal area used for high-resolution vision (the fovea and its surrounding macula), with a relative sparing of peripheral vision. Central vision loss affects quality of life and performance of daily activities such as reading [4], face recognition [5], mobility [6], watching television [7] and is associated with elevated levels of depression [8] and increased mortality [9].

A major challenge in the development of effective treatments for AMD is the lack of sensitive, practical visual function endpoints [10][11] that can be easily measured. People with AMD manifest delayed Dark Adaptation Recovery (DAR) [12][13][14] [15], even in the early stages of the disease [16], and DAR delays become longer with the disease progression [17][18][19]. DAR is therefore an early biomarker for AMD and is an FDA approved method for diagnosing AMD [20]. Measurement of DAR typically utilizes a flash illumination that bleaches rod and cone photoreceptors and the subsequent response recovery serves as a measure of the stress response of retinal metabolism [21][22]. DAR is adversely affected by choroidal thinning [23], morphological changes in retinal pigment epithelial cells [24], and the accumulation of subretinal drusenoid deposits (SDDs) [25][26][27] in AMD. Current DAR tests typically measure psychophysical thresholds for the detection of visual stimuli following bleaching in order to monitor the recovery of sensitivity over time, typically over more than 20 minutes (for review see [28]). Such long DAR testing times are problematic for behavioral testing, especially when both eyes are to be tested. Although rapid (10 minute) protocols have been developed [29], these protocols test a single retinal location which may be useful for classification of AMD patients and control subjects, but they require active participation and attention of the subject. There is therefore an unmet need for a practical, sensitive test that will provide objective, quantitative endpoints for early diagnosis and for monitoring the patient’s response to newly developed treatments.

Visual evoked potentials (VEPs) generated in response to visual stimuli, provide a quantitative, objective measure of the neuro-optical pathways and visual processes that are correlated with behavioral measures of sensitivity [30][31] but do not require sustained attention from the patient. Multi-focal Electroretinogram (ERG) methods have shown deficits in AMD [32] that are correlated with structural defects [33][34], but they require the use of discomforting corneal electrodes. Furthermore, the sensitivity of ERG to scotopic stimulus levels has been shown to be over 2 orders of magnitude worse than either VEP or psychophysical methods [35], so these tests do not serve well as functional measures of DAR. While there have been many studies of VEP in the context of DAR [36][37][35][38], and the use of multifocal VEP has been studied in the context of AMD [39][40][41], we believe that systematic use of this very attractive technique has been hampered by lack of easy-to-administer, affordable instrumentation, and we have not found any previous VEP studies of DAR in AMD, specifically, in the literature.

We have developed a portable, wireless system called NeuroDotVR (illustrated in **Figure 1)** that enables us to measure DAR using an objective paradigm based on transient Visual Evoked Potentials (VEP), which we refer to as Dark Adapted VEP (DAVEP). In this study, we first establish the transition of the photopic (cone photoreceptor-mediated) VEP signal into the scotopic (rod photoreceptor-mediated) response regime, in normally-sighted subjects; wherein, stimulus luminance was stepped from photopic (32 cd/m^2^), through mesopic, down to scotopic levels (10^−3^ cd/m^2^). Then, we evaluate the recovery of the DAVEP response post photobleach (400 cd/m^2^ for 60 s) over a time period of 15 minutes, using a constant scotopic luminance stimulus (5×10^−4^ cd/m^2^) in normally-sighted control subjects, patients with Age-related Macular Degeneration (AMD), and a few patients with diagnoses other than AMD. We developed dichoptic monocular stimuli to test both eyes in two sub-regions of the visual field, “central” (2.6 - 16° eccentricity) and “peripheral” (16 – 33°), independently during the same experimental session. These visual field regions were chosen to provide comparative measures of central (parafoveal macular) and peripheral DAR, because previous studies (that are confirmed in our results below) show, using psychophysical detection thresholds, that AMD patients often exhibit a weak macular response and normal or less impaired peripheral response. We therefore hypothesized that a comparison between these responses provides a robust signal of the disease state in AMD, one that corrects for individual differences in baseline amplitude.

**Figure 1.**
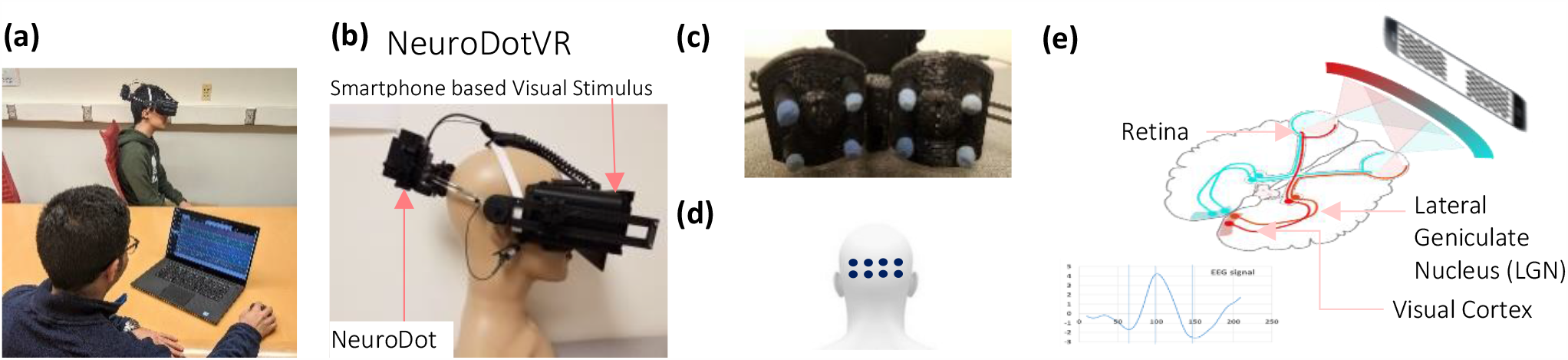
Portable wireless NeuroDotVR system. (a) Operator testing a subject, (b) NeuroDotVR device prototype integrating NeuroDot sensor with visual stimulus headset, (c) Closeup of NeuroDot EEG sensor arrays, (d) Location of electrodes on the scalp over the visual cortex, (e) Human visual pathway and typical photopic visual evoked response measured on occipital scalp, using dichoptic contrast reversing checkerboard stimulus. (Image in part (e) derived from: https://commons.wikimedia.org/wiki/File:Human_visual_pathway.svg)

## Methods

Our DAVEP paradigm makes use of a prototype system for portable neuro-ophthalmic diagnosis, called NeuroDotVR, which combines scalp neuroelectric potential sensors with a smart phone in a portable wireless headset (**Figure 1**). The system records VEPs in response to dichoptic stimuli presented on the smartphone display. [42]

### Neuroelectric Sensing

Each NeuroDot sensor array comprises 4 small-diameter (< 8mm) biopotential electrode pins arranged in a grid with a small spacing of 2 cm (considered to be “ultra” or “very” high-density [43]). The NeuroDotVR system (**Fig. 1a**,**b**,**c**) uses two independently positionable sensor arrays, yielding 8 independent channels of EEG data which, in this study, have been averaged in neighboring groups of 4 electrodes to simulate the three contiguous standard 10-20 System scalp locations *O1, Oz, O2* where the central location, *Oz*, uses the rightmost two channels of the left array and the leftmost two of the right array. Each of the sensor pins connects to its own amplifier channel, which amplifies the potential difference between it and a separate reference electrode, which is clipped to the left ear of the subject. The average potential of the symmetric array (with respect to the remote reference) *V*_*avg*_, is analogous to a single EEG channel at the center of the array, albeit with lower noise. To further simplify the analysis in this study, we take the average over both arrays as the final EEG signal, equivalent to the average over all 8 channels.

The recordings for each EEG channel are sampled at a rate of 1000 samples per second, with a resolution of 24 bits, and a gain factor of 24, and the data is saved, unfiltered, in an binary file format. Before subsequent data processing, each channel is prefiltered using a Stationary Wavelet Transform (SWT) baseline removal [44] with an effective cutoff frequency of 0.5 Hz; we find that this process preserves the time domain characteristics of EEG stimulus responses while greatly reducing small motion artifacts and electrode polarization artifacts that might otherwise create longer duration transient distortions (impulse/step response of filter) using standard digital filtering techniques. Any additional filtering steps will be described in the “Results” sections below.

### Headmount

Using 3D printing technologies and off-the-shelf hardware, we fabricated a purpose-built headset that is compatible with the Google Daydream VR platform. Our custom design targets full light-tightness, minimal light leakage between apertures, and incorporates mounting mechanisms for the electrode sensor arrays. The headset also features a concealed mount for a phototransistor sensor that is used for accurate timing of stimulus onset and reversal. A custom foam padding was molded for comfortable use using silicone replica casting from a 3D printed model.

### Visual Stimuli

Pattern reversal dartboard (polar checkerboard geometry) “test” stimuli were applied as probes of DAR over 15 minutes after exposure to a 400 cd/m^2^, 60 s white “photobleach” stimulus. The display device was the OLED screen on the Google Pixel 2 XL smartphone, used with a custom VR head-mounted-display having optics similar to the Google VR Daydream View (first version, 2016). On LED displays, the black pixels emit no light, so contrast is effectively close to the maximum that subjects can perceive, assuming minimal reflected light in the viewer. The stimuli patterns were generated as high-resolution pixel maps (2048×2048) using custom Python programs, then were loaded into a custom app using the Google VR Android SDK with OpenGLES2 texture rendering. The dartboard was divided into two regions: the central region, labeled “Macular Anulus” (or “MA”), including all of the macula but excluding the fovea (2.6 - 16°) and “Near Peripheral Anulus’’ (or “PA”) region (16 – 33° eccentricity) of each eye as annular dartboard patterns that scale radially according to the rule of cortical magnification [45], where each check radial length *R* at eccentricity *E* (in degrees of visual angle) scales by a factor of *R*(*E*)*/R*(0) = 0.27*E* +1, and the base checksize R(0) = 1° of visual field subtended. **(Figure 2)**

**Figure 2.**
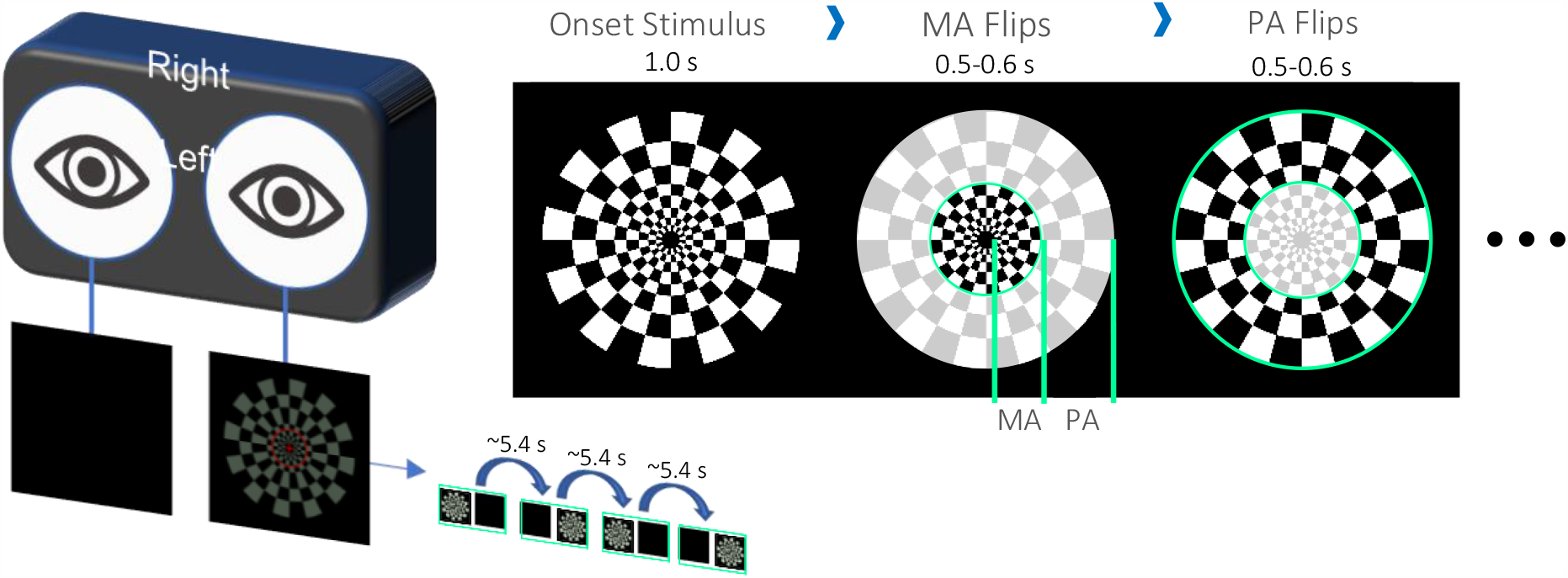
Monocular stimuli for DAVEP paradigm: log polar dartboard pattern is divided into the “Macular Anulus” (MA, 2.6-16° ecc.) and “Peripheral Anulus” (PA, 16-33° ecc.) visual field regions whose check pattern reverses independently – actual stimuli are rendered as solid green pixels. In the VR viewer, stimuli are presented monocularly (to each eye with the opposite eye completely dark) with a brighter red fixation ring and cross. Under scotopic conditions, all stimuli appear grayish. After an unmeasured “onset” period of 1.0s, the pattern reverses 3 times for each visual field region with a step of 0.5-0.6 s (randomized); the sequence lasts about 5.4 s before switching to the other eye. See the text for an exact description of the sequence.

The stimuli were solid green colored (wavelength 528 ± 30 nm, single channel on RGB display, closest to the rod photoreceptor sensivity centroid of 498 nm), although they appear virtually colorless at scotopic intensity levels. The foveal region of 0-2.6° was excluded in order to minimize the contribution of cone photoreceptors which have highest density here. The choice of extending the “MA” region to 16° eccentricity beyond the physiologically defined macula at 6° was made in order to ensure that a roughly equitable amount of rod photoreceptors would be stimulated in each region (rod density increases from zero outside the fovea to a peak at 18°, but falls off more slowly in the periphery) [46]. A neutral density filter of 8 stops (log base 2 units) was employed to achieve scotopic luminances of around 10^−3^ cd/m^2^ which was not possible using *only* the native display range; this stimulus level was chosen in the upper scotopic regime to limit the expected response amplitude recovery times for normal subjects below the 15 min experiment durations (see next section). A mid-mesopic intensity red fixation cross, spanning the foveal region, and a thin ring at 10° eccentricity were overlaid on the stimuli in order to help subjects, especially those with central vision loss, maintain fixation at the center.

### DAVEP Recovery Paradigm

In the DAVEP recovery paradigm, following the photobleaching stimulus, a continual series of scotopic luminance level test pattern reversals is presented according to the fixed sequence, where S is starting image (1.0 s onset, not measured), and M and P are macular and peripheral region reversals, respectively (at an interval of 0.5 s plus a random jitter from 0 to 0.1 s): S, M, P, M, P, P, M, P, M. In the monocular version of the paradigm, this sequence is cycled between the left L, then the right R eye – lasting ∼5.4 s in each eye, where the opposite eye is in complete darkness. The NeuroDotVR system records accurate start of video frame events using a phototransistor detector to monitor a hidden patch of the screen used exclusively for sending this synchronous signal. These stimulus event markers are used in subsequent data processing steps to average the neuroelectric responses at the precise start time of these stimulus presentations. Epochs are defined from the start of the stimuli up to 600 ms (by when the response is expected to have already decayed below the noise floor) and grouped separately by L or R eye and M or P region (S stimuli onsets are not processed). The average over the LM, LP, RM, RP trials is performed in a centered sliding window that typically includes 121 trials (containing responses spread over a period of ∼3.6 min), with a step size of 7 trials (∼15 s) creating a smoothed response profile over the course of the experiment (see 3D wireframe plots in **Figure 5**). Using an unsupervised machine learning outlier rejection algorithm (Scikit-Learn’s “Isolation Forest” model [47]), trials with outlier variances (e.g. arising from movement artifacts) are excluded from the windowed averaged – typically a small percentage (up to 10%).

### Subject Pool

All studies were approved by the Northeastern University Institutional Review Board and were performed in accordance with the Declaration of Helsinki. All subjects were either referred by their clinical ophthalmologist or were recruited from personal contacts at Northeastern University. Subjects were required to be greater than 18 years of age and had to sign an informed consent document after their role in the study was explained. Sex was not used as a condition for selection.

Because of the VR capabilities of our display system, we required subjects to fill out a simulator sickness questionnaire; however, the static nature of our stimuli was not expected to induce motion sickness symptoms, and this was borne out in the results (see below section “Subject Tolerance”). Four of the authors served as control subjects (CV, SMAB, PB and SS), who have extensive experience participating in EEG and/or psychophysical vision testing paradigms.

The dichoptic (monocular) DAVEP recovery experiments were tested in:

1. (13 × 2) = 26 eyes with AMD. The AMD subjects age range was 71 to 89;
2. (5 × 2) + (3 × 1) = 13 normal eyes. The age range was 21 to 73;
3. (3 × 1) = 3 eyes with other conditions. The age range in this group was 51 to 73.

(See **Table 2** at the end of the Results & Discussion section for a detailed overview of subject characteristics.)

### Test Administration

Most clinical subjects were new to EEG/VEP, but were instructed to minimize body movements, eye closures, and talking during the recording; however, they were explicitly permitted to blink and shift posture as needed for comfort. No drugs for pupil dilation were administered and no artificial restricted pupils were used for any of the reported tests. After fitting the head mounted display and neuroelectric sensors to the subject, the experimenters applied a skin-safe saline electrolyte and disinfectant solution with gentle swabbing directly to the area around the electrode/scalp contact, while monitoring impedances of the EEG sensing channels and ensuring that they were below 100 kOhms – which we have previously shown to be adequate for high quality measurements using the NeuroDot system [48]. No abrasive preparation compounds nor conductive pastes or gels were applied to the scalp. Any bulky jewelry or headwear that could interfere with electrodes or the mounting hardware was removed prior to experimentation. Subjects were asked to remove any spectacles, *which is not typical for photopic VEP experiments*, but we had previously determined that the DAVEP stimuli-responses are largely unaffected by corrective eyewear. The tests were minimally discomforting but demanded subject alertness for an extended period of time (about 15 min) in the presence of repetitive visual displays at low light levels; subject boredom and microsleeps were, therefore, a concern, so we encouraged subjects to listen to their favorite music or a podcast for the duration of testing.

## Results & Discussion

### Subject Tolerance

There were no adverse events during testing. To monitor subject tolerance to the test, we administered a simulator sickness questionnaire that is a modified version of the Pensecola Motion Sickness Questionnaire [49] (provided by Prof. Yingzi Lin of the department of Mechanical & Industrial Engineering at Northeastern University). The form consisted of 24 questions with a 4-point rating scale (none/slight/moderate/severe), before and after the test. For most of the questions and most of the subjects, there were no changes attributable to the test. We’ve listed in **Table 1** below only the subset of those symptoms that did differ at the end of the test, which shows that it was generally well-tolerated.

**Table 1.** Assessment of symptoms of adverse events during testing

| Symptoms | No. of Subjects | Change |
| --- | --- | --- |
| General discomfort | 3 | None to Slight |
| Aware of breathing | 3 | None to Slight |
| Drowsiness | 3 | None to Slight |
| Salivation decreased | 2 | None to Slight |
| Fatigue | 1 | None to Slight |
| Boredom | 1 | None to Slight |

### Evolution of VEP from Photopic to Scotopic Stimulus Regimes

In order to better understand the relationship between the lower photopic, mesopic, and upper scotopic level responses to our green dartboard stimuli, we undertook a pilot study that stepped down luminance (by −0.5 log units every 2.5 minutes) over the range 32 to 0.001 cd/m^2^ over the course of 25 minutes on two normally-sighted subjects (authors CV and SMAB), who have experience sitting for long VEP paradigms. Subjects had preadapted to dark conditions for at least 30 mins prior to recording. The Macular Anulus (MA) and Peripheral Anulus (PA) visual field regions were tested alternately in only the left eye at an interval of 1.0-1.1 seconds (randomized). EEG signals were processed with the 0.5 Hz cutoff baseline removal (highpass) filter described in “Methods – Nueroeletric Sensing” and further filtered with a 30 Hz cutoff Bessel lowpass Infinite Impulse Response (IIR) digital filter. An epoch length of 600 ms post pattern reversal event was chosen for analyzing and displaying VEP signals, since we observed no measurable changes after this period. A sliding window of 61 trials with step of 3 trials was used to create a smoothed VEP amplitude over the time course of the experiment by averaging epochs for the same visual field location. The data from the macular and peripheral regions of one normally-sighted subject is shown in **Figure 3**.

**Figure 3.**
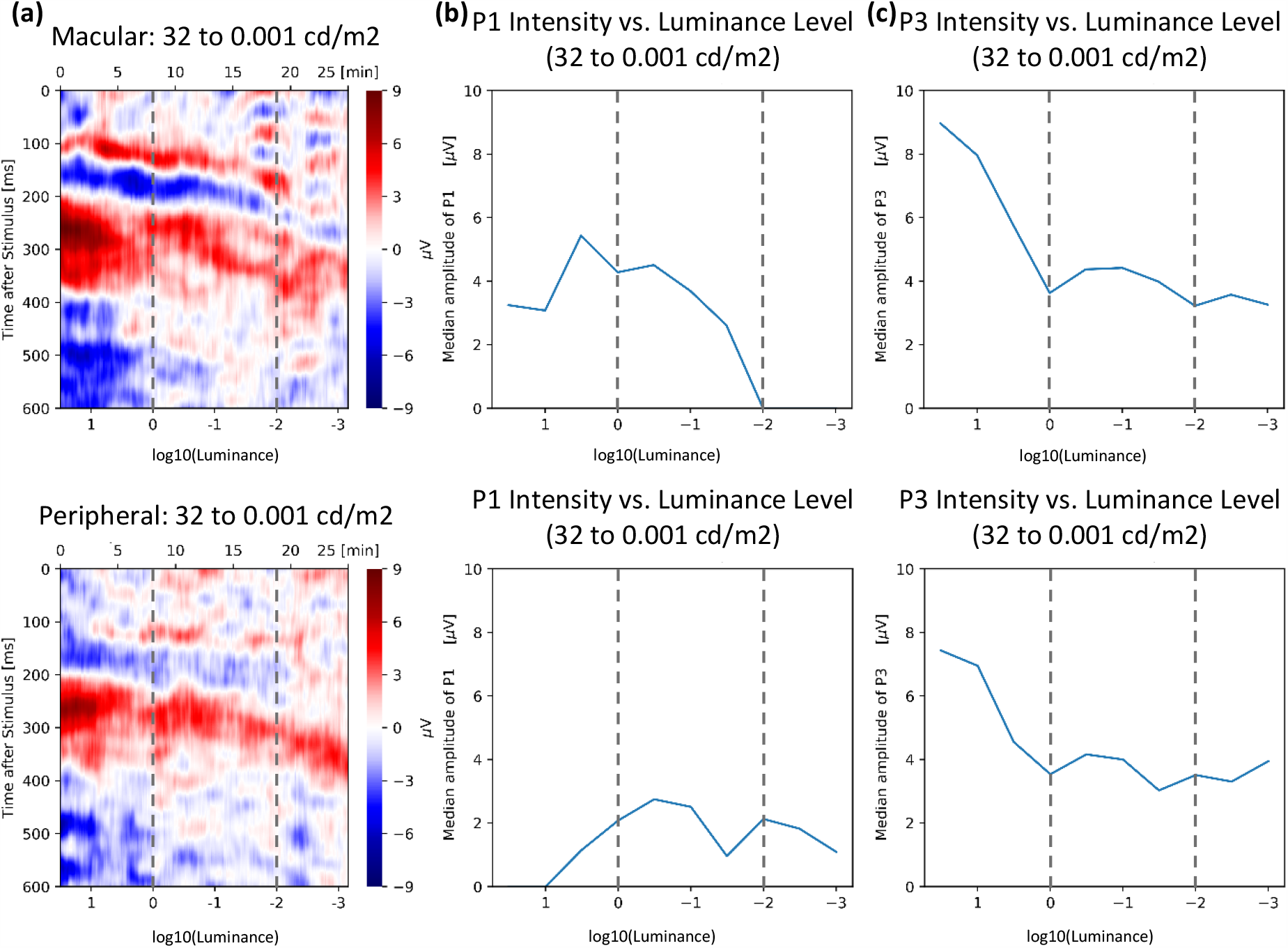
**(a)** The heatmap plots show VEP response amplitude for one normal observer in MA (top) and PA (bottom) visual field regions using a color scale over the range +/-9 microvolts, where vertical axis shows time (latency) after the start of the stimulus trial (up to 600 ms) and the horizontal axis spans the duration of the experiment (25 mins) with the log brightness scale at the bottom. The following plots show the luminance dependence on the amplitude of the **(b)** positive peak near 100ms, P1 **(c)** and the positive peak near 300 ms, P3 or P3^DA^ (see text). For **all plots**, the dashed lines approximately separate the stimulus intensities, from left to right, into regimes of active photoreceptors: “photopic” (only cones), “mesopic” (cones & rods), and “scotopic” (only rods).

From these studies we drew the following conclusions: **(1)** the typical P1 response (positive peak at approximately 100 ms) seen in many photopic VEP paradigms, but here only obvious for the cone-rich macular region, decreases rapidly over the mesopic luminance range (where both cones and rods are active, 0 to -2 log units) and is not present in the scotopic response (where only rods are active, below -2 log units); note that the photopic response may have been further reduced from standard levels by the exclusion of the foveal region from stimulation and the use of monochromatic green pixels; **(2)** the positive peaks in the range 200-300 ms, seen at the initial photopic stimulus levels (often referred to as P2 & P3 or just P300 [50], [51]), decrease even more rapidly; in its place, a new positive component (or set of components) around 200-300ms, here labeled P2^DA^ & P3^DA^ (superscripts denotes appearance under dark-adapted conditions) emerges in the mid-mesopic regime and persists down to scotopic levels (-0.5 to -3 log units) but shifts to later latencies by approximately 100 ms or more around our DAVEP testing level (5×10^−4^ cd/m^2^) (see **Figure 4** for a schematic description of VEP peak labels); the peripheral P2^DA^ & P3^DA^ response is similar to that of the macular, but arguably a bit simpler and less contaminated by Alpha waves (see next point); **(3)** in some subjects, low amplitude residual noise peaks in the Alpha band (8-12Hz) are visible but are not strongly time-locked to the stimulus presentation; this Alpha band contamination has been noted in previous scotopic level VEP studies [37] – care must be taken to avoid false-positive response measurements where Alpha band amplitudes may dominate even after time-locked stimulus averaging.

**Figure 4.**
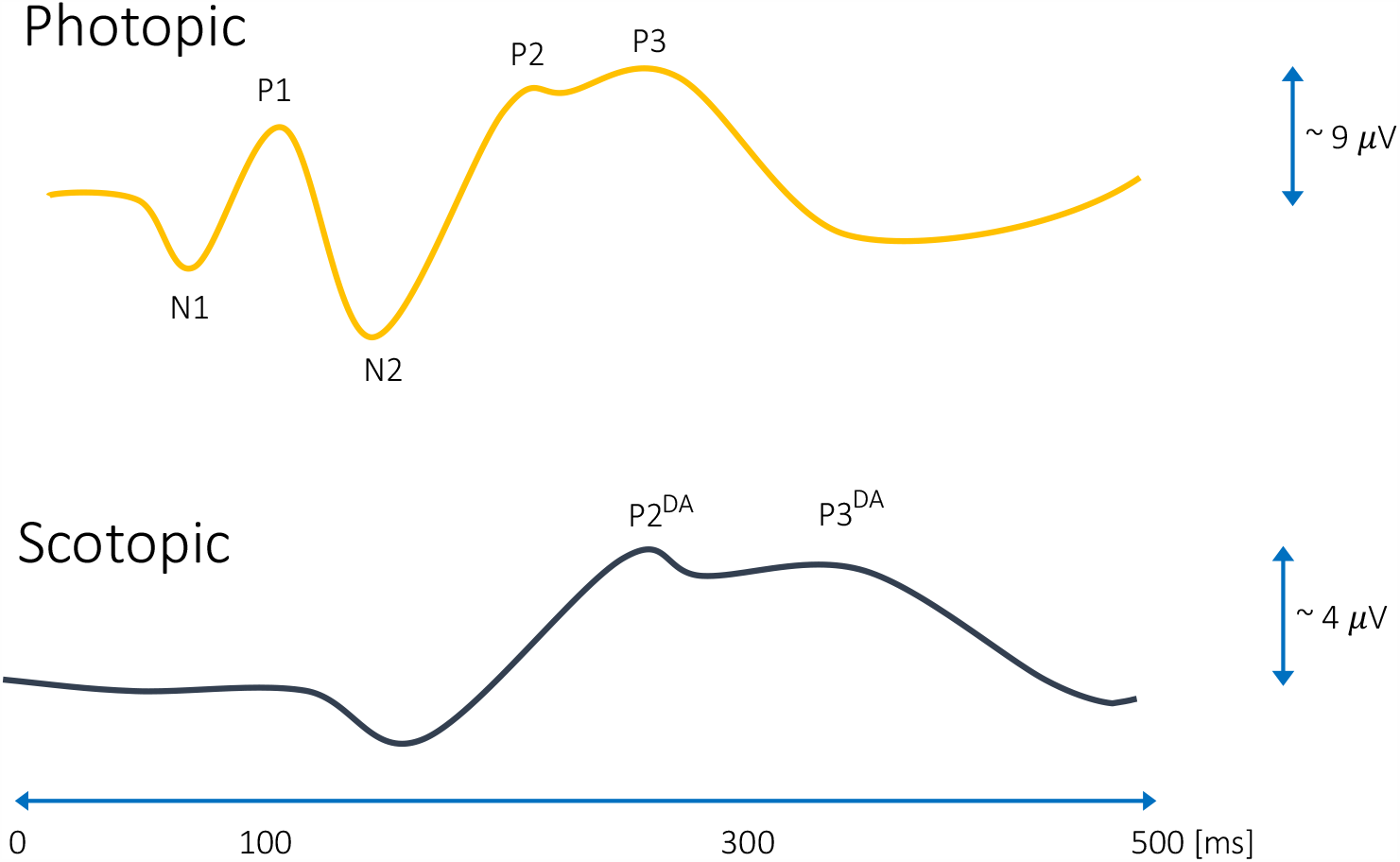
Schematic of the typical VEP components seen in this study for dartboard pattern reversal stimuli in normally sighted subjects. At photopic levels the response is similar to classic pattern reversal VEP, featuring a N1 (∼75 ms), P1 (∼100 ms), N2 (∼150 ms), P2(∼225 ms, not always present/distinct), P3(centered around 250-275 ms); however, the N1 & P1 response is of lower amplitude (in MA region) or not detectable (PA region) since monochromatic green light is used and the foveal field is not stimulated. At scotopic levels, under dark adaptation (DA), the largest amplitude components are labeled P2^DA^ (around 275ms, narrower, not always present/distinct, varies by subject) and P3^DA^ (centered around 350-400 ms, broader, more reliably detected across subjects).

Previous studies of scotopic *full-field flash* VEPs [52][53][54] contain analyses of positive peaks with latencies around 200 ms but ignore the lower amplitude positive peaks around 300ms; however, we caution against a direct comparison since those stimulus types are much different than our *restricted eccentricity pattern reversal* VEPs. The closest match to our stimulus conditions in the literature [55], scotopic pattern-reversal in restricted central and near peripheral visual fields, is presented as a brief study on VEP scalp distribution in normal subjects; we note here a similarity between this previous literature study’s figure 3 (right side) and our study’s P3^DA^ responses.

We tentatively interpret these results to mean that while the typical photopic VEP peak amplitudes are highly sensitive to diminishing cone photoreceptor input and, possibly, to suppressive rod-cone interactions [56][57][58], the emergent P2^DA^ & P3^DA^ response serves as a marker of *cognitive visual awareness* under lower mesopic and scotopic conditions; therefore, this response is contingent on rod photoreceptor stimulation and subsequent pathway latencies, but the amplitude is relatively insensitive to stimulus luminance under full DA, at least in this tested range. Given the high latency and stable amplitude, this VEP signal might be related to higher level cortical processing than the assumed early V1 cortex source of the photopic P1, possibly in the same class as other P300 event related potentials (ERPs) such as that which is manipulated under “visual odd-ball” paradigms [59]. Our further results focus on these scotopic P2^DA^ & P3^DA^components (and, potentially, a complex of positive and negative response components between latencies of 175-450 ms, seen in other subject data) as a marker for DA recovery after photobleach (adaptation to bright conditions) in normal subjects and AMD patients.

### Post Photobleach Recovery of DAVEP Response Shows Deficits in AMD Subjects

In the DAVEP Recovery experiment, subjects previewed the stimuli at a mesopic luminance level for approximately 2 mins, followed by a 60 s 400 cd/m^2^ white photobleach conditioning (covering the full tested visual field); then, subsequently, the stimulus was switched to a scotopic luminance level (5×10^−4^ cd/m^2^), while recording their neuroelectric responses – principally the amplitude of P2^DA^ & P3^DA^ components described above – for 15 minutes. The EEG signals were processed to derive stimulus time-locked DAVEP signals separately for each eye and each visual field region. The 8 EEG channels were first prefiltered using a Stationary Wavelet Transform (SWT) baseline removal [44] with an effective cutoff frequency of 0.5 Hz; we find that this process greatly reduces small motion artifacts and electrode polarization artifacts, while preserving the principle shape of the VEP time domain signal. Alpha band noise, which may cause false positive signal, was present in many subject’s DAVEP response, especially immediately after the photobleach; since the P2^DA^ & P3^DA^ response components have relatively lower frequency content (is more slowly varying), we designed an SWT lowpass filter with a sharp cutoff at 7Hz, right below the Alpha band. Although such drastic filtering may highly smooth the time domain shape of the DAVEP, the recovery metric (see section below) that we are using in this study is mostly an integrated measure of amplitude within a broad response time window (175-450 ms), so it would be only marginally distorted. Finally, the 8 channels were averaged together and a sliding window of 121 trials (containing responses spread over a period of ∼3.6 min) with a step of 7 trials (∼15 s) was used to create a smoothed VEP amplitude over the time course of the experiment by averaging epochs (excepting those marked as containing outlier variance artifacts, see section “Methods: DAVEP Recovery Paradigm”) for the same eye/visual field location. The same data processing steps were applied to all subjects using a custom fully automated Scientific Python software framework; although, the artifact epoch rejection step is adaptively computed based on a subject’s global trial statistics.

The VEP response over the DA recovery period from the macular anulus (MA) and peripheral anulus (PA) visual field regions of one normally-sighted control subject and one AMD subject is shown in **Figure 5**. From the aggregate DAVEP data of all subjects, we drew the following conclusions: typically, **(1)** the recovery of the P2^DA^ & P3^DA^ response components is rapid for a healthy subject in both the macular and peripheral regions, while little to no recovery can be seen, specifically, in the macular region of the AMD subjects; **(2)** there is a noticeable difference between the macular and peripheral response of most AMD subjects, with the MA amplitudes being weaker and recovering later than the PA (which was often indistinguishable from a normal response).

**Figure 5.**
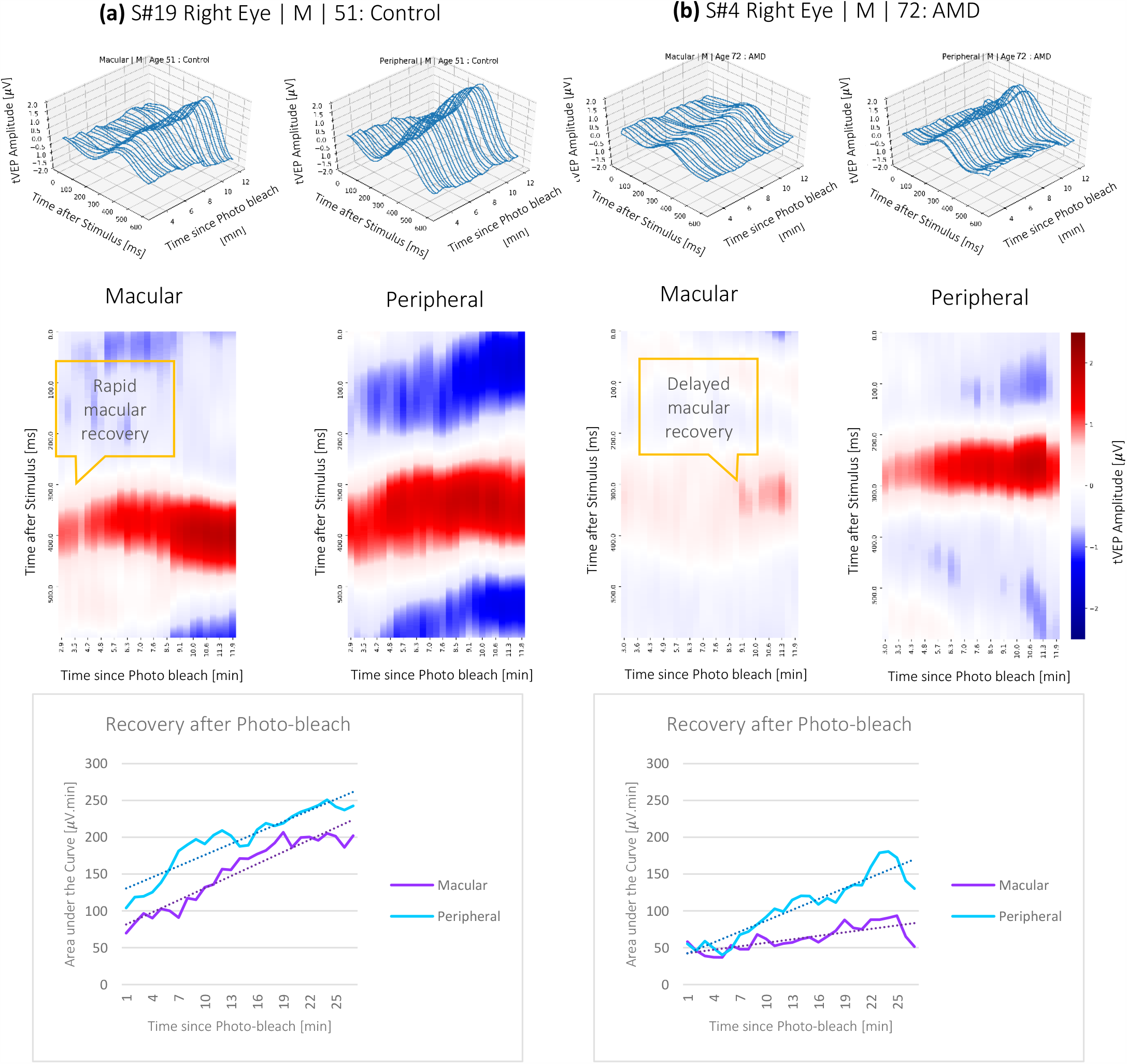
The plots show the recovery of the vision for the right eye of **(a)** Control subject, and **(b)** AMD subject in the macular and peripheral regions. The 3D wireframe plots (top row) show the windowed DAVEP response wave (X axis is time after stimulus start in ms and Z axis is the voltage amplitude in microvolts) as it recovers following the initial photobleach condition (minutes along Y axis). Note that, in the heatmap plots (middle row), the vertical axis shows the response wave timing (in ms after stimulus start), the color axis shows the response polarity and amplitude (in microvolts), and the horizontal axis shows the time passed after the photo bleach (in minutes). The bottom row displays the area under the curve (µV.min) for the component between 175 to 450 (ms) on the Y axis vs. the time passed after the photo-bleach (min) and indicates the recovery of the vision. Notice the steep recovery rate in the macular region of the control subject vs. the slow rate of recovery for the AMD subject.

The exact characteristics of this emergent response under dark adaption is presently not well understood: some subjects show only an early narrower peak centered around 275 ms, distinguished by the label P2^DA^, many show a broader peak between 300-400 ms, labelled as P3^DA^; some show a mixture of responses that may even change with state of arousal or with the time post photobleach and may do so differently between the two tested visual field regions, MA & PA; some, especially AMD subjects, show a greatly diminished response particularly in the MA visual field region but may have a normal level response in the PA region.

### DAVEP1 Score Reliably Classifies AMD Subjects

Based on the findings above, we developed a metric that objectively characterizes DAR using the DAVEP signal response over the finite 15 minutes post photobleach time course. At each time window over the trials, a K-means clustering algorithm [47] is used to dynamically find the center of the prominent response cluster (within the latency range of 175-450 ms) and extract the amplitudes spanning latencies in the “response window” within +/-75 ms of the center. This automated response analysis is intuitively similar to the choices an expert technician might make. These responses are then used to compute the “DAVEP1 score” across the recovery time period: in order to correct for substantial variations in DAVEP response amplitude across subjects, the amplitude values of each visual field region and each eye are normalized *within the subject* by the maximum value obtained *across all locations*; then, the score is calculated as the *spectral norm* for the matrix containing these smoothed, normalized amplitudes within the response window:

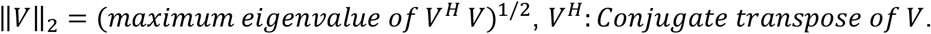

The score is indicative of the continuity and normalized intensity of the responses obtained in that location, i.e. an early recovery with a sustained amplitude close to 1.0 maximizes the score. The ratio of the macular to peripheral score is also calculated for each eye, which emphasizes the functional disparity between these visual field regions. A very low score in the macular region compared to a high peripheral score is potentially indicative of AMD (a ratio of macular to peripheral score of lower than 0.6).

Based on the available data (see **Figure 6** & **Table 2**), we found that macular scores below 25 (red area in the plot) are indicative of deficits in DAR and is associated with subjects who have problems with their macular vision, and scores of above 30 (green area in the plot) are indicative of a healthy subject. Macular scores between 25 and 30 put the classification into the “watch” region (yellow area in the plot), where a closer look at the data is necessary: if the ratio of macular to peripheral score in this region is higher than 0.6, it still points to a healthy performance but any subject’s eye with a macular to peripheral score ratio lower than this threshold is classified as potentially having AMD. Finally, the algorithm marks a subject’s eyes with a peripheral score below 25 *and* a macular score above 30 as “abnormal” (gray area of the plot); these “abnormal” results are withheld from classification – possibly due to other confounding vision issues or, perhaps, problems with fixation during the test. For example, it is common for AMD patients use a preferred retinal locus (PRL) [60] in the peripheral field for reading and other tasks. It is recommended to examine the heatmap plots of the responses and, if available, other diagnostics such as acuity to make a more detailed justification in these cases.

**Table 2.**
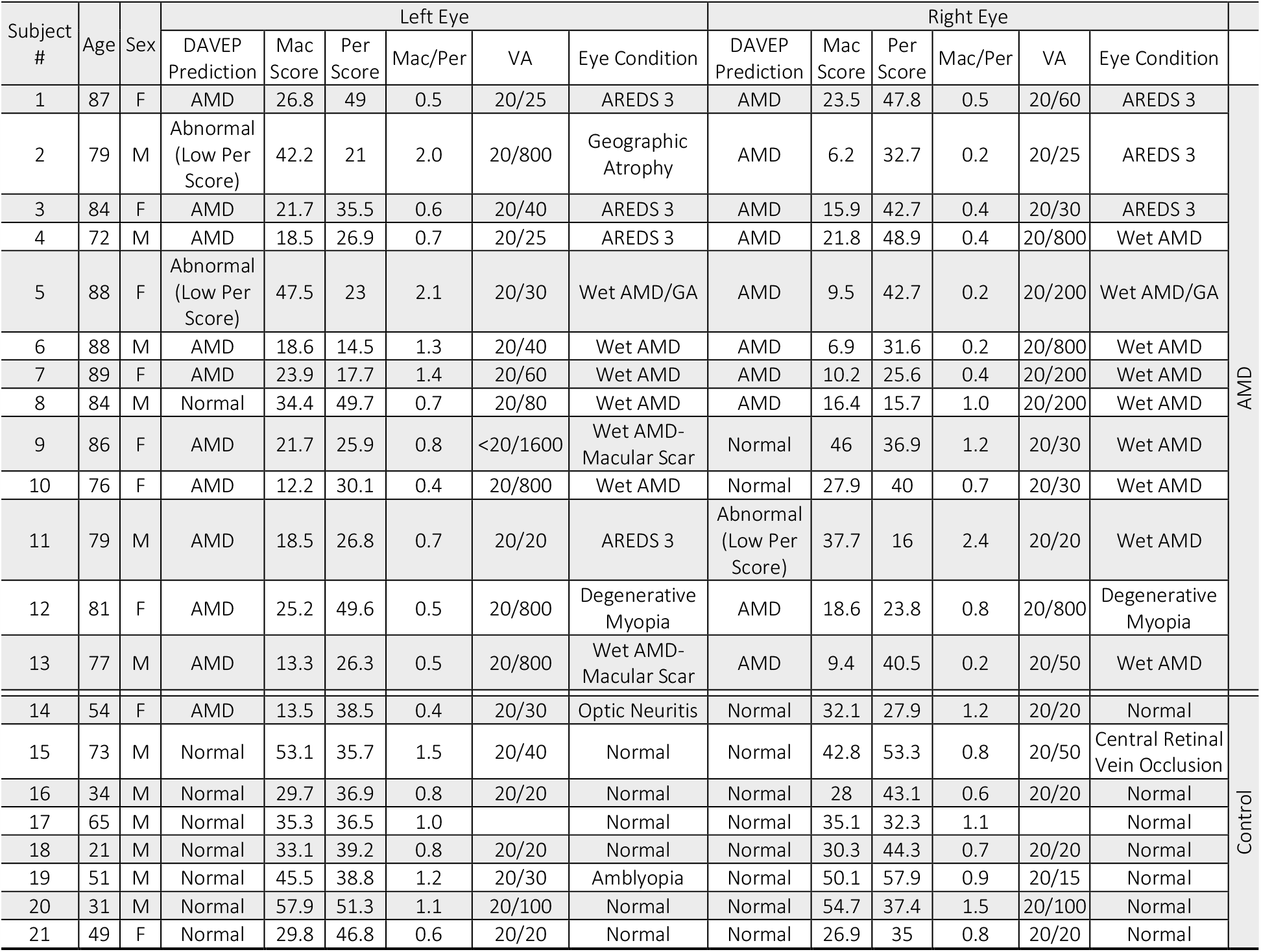
Details of DAVEP1 scores among subjects

**Figure 6.**
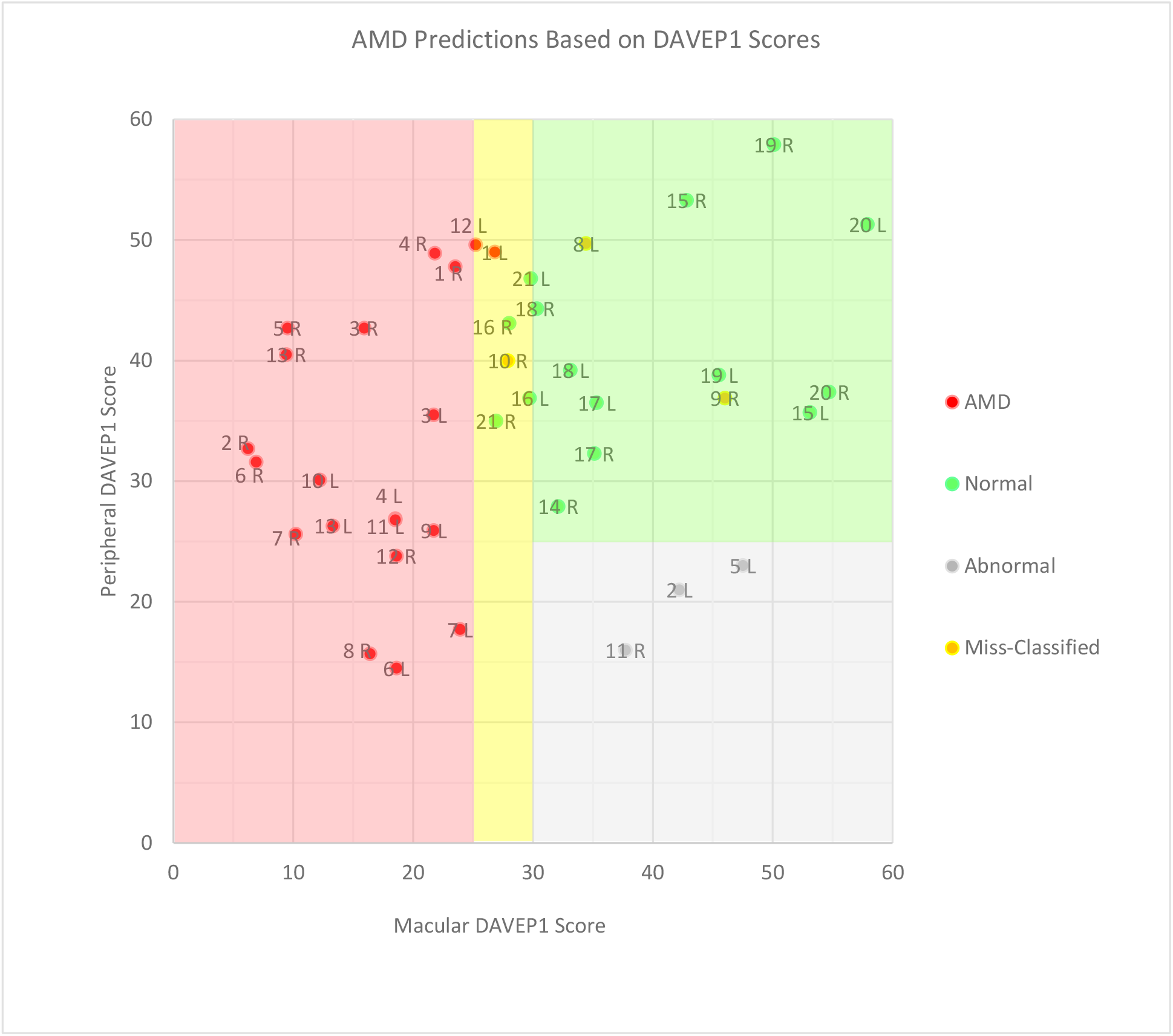
The chart shows the distribution of the macular vs. peripheral DAVEP1 scores for all subjects and the table shows the details of the classification for left and right eyes based on these scores vs. the clinical diagnosis of the subject. The yellow area marks “watch” region where distinguishing between AMD and non-AMD subjects is based on the macular to peripheral DAVEP1 score ratio (< 0.6 indicates AMD). The color of the points displays the outcome of our algorithm predictions according to the legend. The labels show subject number and whether it is the left or right eye. It is expected to have comparable scores for the two visual field regions between both eyes in a healthy subject. Based on the prediction criteria, the plot can be divided into four regions: 1. Top left quarter (in the red area) where we have the most obvious AMD cases with low macular and high peripheral scores; 2. Bottom left corner (also in red area) where the subjects have low scores in both macular and peripheral regions – perhaps indicating advanced disease status; 3. Top right quarter (green area) which is the region for both having high score, presumably indicating healthy DAR; 4. Bottom right quarter (gray area) marks a few outlying cases where the eyes have low peripheral and high macular scores (these are marked by our algorithm as “abnormal” and requiring further diagnosis).

The DAVEP1 score successfully classified 90% (36/40) of subject eyes. AMD was correctly classified in 87% (20/23) of eyes, with 13% (3/23) false negative results and 3 eyes having been withheld from classification because of “abnormal” performance as described above. In all 3 false negative cases, the misclassification was for the eye with higher visual acuity and AMD was correctly classified in the patient’s weaker eye. Likewise, in the 3 “abnormal” cases, the fellow eye was classified as AMD positive. Therefore, the algorithm correctly classified the presence of AMD in at least one eye of 100% of AMD patients. 94% of eyes (15/16) of control subjects were correctly classified, with 6% (1/16) false positives. The single false positive result happened in the case of a patient with optic neuritis and cortical age-related cataract, but no AMD.

The use of a peripheral PRL to view all stimuli may, conceivably, lead to the “abnormal” inversion of peripheral and macular scores detected in our study. It was the case that one of the AMD patients (#5) who received an “abnormal” result in one eye (the one with higher visual acuity), claimed to have not seen the stimuli (including the brighter, static fixation aids) at all, even though a strong response was detected; these factors may be indicative of visual suppression [61]. Under the monocular conditions of our experiment, both eyes are left open while the un-stimulated eye is kept in darkness until it is stimulated in-turn (switching approximately every 5 seconds) – this suggests, perhaps, a follow-up assessing just the responding eye in complete isolation may be helpful. Another of the “abnormal” results was from a subject (#11) that, despite, having an AMD diagnosis, had 20/20 visual acuity in both eyes, and was noted to have been pupil-dilated in a clinical check-up immediately before participating in our study; it is conceivable that a peripheral PRL, especially in a dilated eye, could cause a large false response to be attributed to macular region trials which were assumed to be fixated at the center. In future developments of DAVEP paradigms, the integration of eye-tracking cameras might help to disambiguate abnormal results and further improve classification results across the board through rejection of poorly fixated trials.

While the DAVEP1 score can be used to classify most subjects reliably, it is not a complete representation of the data recorded within the DAVEP Recovery paradigm. For example, each time window is given equal weight, even though early windows are more informative for predicting healthy function and later windows are more informative for assessing the degree of DAR impairment. When subjects do show early recovery, there is typically a phase of monotonic amplitude increase which may be followed by a lull in response amplitudes, possibly due to higher order cognitive adaptation to the repetitive signal (perhaps hinting at the potential brain sources of the P2^DA^ & P3^DA^ complex in later visual cortices or in parietal lobes). On the contrary, for subjects who have significant delays in DAR, the earliest epochs contain only noise which may contribute false signal to the DAVEP1 score (this is the main reason why we chose to filter out all frequencies at and above Alpha band, a major noise source in many subjects). Furthermore, for simplicity in assessing the paradigm, only the responses for the average of the 8 spatially distributed EEG channels (covering scalp locations O1, Oz, O2) were analyzed. While this spatially averaged montage reduces noise and is appropriate since the EEG potentials are largely homogenous over these locations, this data reduction strategy does not take full advantage of the neuroelectric information contained in the recordings.

The DAVEP1 score is a first order approach to AMD patient classification, which is expected to generalize reasonably well to patients of unknown disease status. In addition, much more information can be extracted from the existing DAVEP Recovery paradigm recordings, specifically regarding: the time-order dependence of recovering VEP signal properties; the spatial distribution of EEG signal variations over the covered O1, Oz, O2 scalp locations; and the change in other spectral characteristics, such as Alpha band power. As the feature sets over the signals expand, the manual analysis of their interdependencies becomes increasingly infeasible. We see the future potential of more automated machine learning approaches to our data analysis tasks and for other complex electrophysiology paradigms.

## Conclusions

The results of this study confirm that using VEP during DA Recovery for AMD symptom testing, implemented on the portable NeuroDotVR platform, is well-tolerated and can be successfully deployed in a busy clinic on naïve patients with various levels of visual impairment. The DAVEP test, during which the subject can listen to audio entertainment, is comfortable. The DAVEP test paradigm is completely objective and does not require subject response. This technique can measure DAR at 2 locations in 2 eyes in 15 minutes or less, unlike the existing psychophysical tests for DAR based diagnostics which require active subject participation and test individual eyes in a single location.

This study has revealed some new aspects of VEP under dark-adaptation that have not previously been reported. Surprisingly, the intensity dependent data clearly show the disappearance of the commonly observed N1, P1 complex for times < 200 ms at lower luminance and the emergence (or persistence) of components with higher latency. We have found no explicit literature references to the response component we have here labeled P3^DA^, though peaks in this range are often visible in scotopic VEP study data and we note the similarities to the cognitive P300 response seen in many ERP paradigms (mainly high latency and amplitude robustness to stimulus property changes). Although its source remains to be identified, we have shown that the P2^DA^ & P3^DA^ response complex can serve as a robust, objective biomarker of visual awareness under DA.

Our results show that, following a photobleach conditioning (photopic brightness adaptation), the initially suppressed responses for healthy subjects gain amplitude rapidly (typically peaking by 8 min or sooner) but may show a moderate slowing down of recovery with age; in the cases of AMD subjects, the DAR is significantly delayed to later times after the photobleach in the macular annular region, and in some cases in the peripheral annular region as well but to a lesser extent. We developed DAVEP1 scores, a simple metric for DAR, that combines the signal recovery amplitudes and times. Using this score we successfully identified all 100% of AMD subjects and classified 90% of subject eyes correctly. Deficits in DAR in patients with AMD can be identified with this objective VEP based system using the DAVEP1 metric; it is a promising new objective biomarker for this disease that can be implemented in the clinic or other locations. Further studies are suggested to validate the DAVEP approach in a larger cohort of subjects.

## Data Availability

De-identified data sets may be made available upon request to the corresponding author via email.

## Acknowledgements

Work partially supported by HHS-1R41AG057250-01.

Thanks to Dr. Elias Reichel of Tufts Medical Center for invaluable discussions on AMD symptomology.

